# Silicone Induced Granuloma of Breast Implant Capsule (SIGBIC) diagnosis: Breast Magnetic Resonance (BMR) ability to detect silicone bleeding

**DOI:** 10.1101/2020.01.15.20017350

**Authors:** Eduardo de Faria Castro Fleury

**Author notes:** CORRESPONDENCE: Rua Maestro Chiaffarelli, 409 – Jardim Paulista – São Paulo/Brasil. ZIP: 01432-030. DISCLOSURES: There is no grant founding/financial support for this manuscript. Nome of the data presented in this study have been published previously. The authors declare that this study has no conflicts of interest with previously published studies. The authors declare that no conflicts of interest are associated with this research and that this study was not funded by any agency. The study was approved by the institutional ethical committee. Inform consent was obtained from all of the participants. Study protocol: Plataforma Brasil **CAAE:** 77215317.0.0000.0072. Silicone Induced Granuloma of Breast Implant Capsule (SIGBIC) diagnosis: Breast Magnetic Resonance (BMR) ability to detect silicone bleeding.

## Abstract

**Objective:** To evaluate the ability of BMRI to detect silicone gel bleeding in a prospective observational study including consecutive patients referred for BMRI scan. Methods: From January 2017 to March 2018, patients referred for BMRI were evaluated in a prospective observational study. Patients who had breast implants were included. BMRI recorded 9 findings according to BI-RADS lexicon and SIGBIC findings, considered equivocal features to detect gel bleeding (GB). Three new original imaging features were added for SIGBIC diagnosis: black drop signal; T2* hypersignal mass; and delayed contrast enhancement, considered as irrevocable signs. The presence of silicone corpuscle was confirmed by percutaneous biopsy or surgical capsulectomy. Accuracy of BMRI SIGBIC findings to predict GB was determined. We also used univariate analysis for the equivocal features for GB diagnosis. The Backward method was applied for a multivariate Logistic Regression model for the equivocal features. Results: SIGBIC was diagnosed in 208 patients and GB was histologically confirmed in all cases. No false positive results were observed. The most important imaging equivocal feature associated with GB was capsular contracture. In order of prevalence, the main equivocal BMRI features associated to GB with statistically significance (P < =0.001) were as follows: 1.water droplets (OR=2.8; 95%CI 1.8-4.4); 2.enlarged intramammary lymph node (OR=3.1; 95%CI 1.5-6.1); 3.pericapsular edema (OR=5.0; 95%CI 2.3-11.1); and 4.intracapsular seroma (OR=2.4; 95%CI 1.4-4.1).

**Conclusion:** SIGBIC diagnosis has high sensitivity to predict GB by the 3 irrevocable BMRI features described by the authors. We suppose GB is underdiagnosed in clinical practice by BI-RADS features.

## INTRODUCTION

Over the last 2 years, an increasing number of studies have reported complications related to breast silicone implants (1–4). These complications are commonly associated with the incidence of breast implant-associated anaplastic large cell lymphoma (BIA-ALCL) (5–12); however, the onset of this pathology and its trigger point are yet to be elucidated.

Interestingly, many of these changes have been reported despite the macroscopic integrity of breast implants. The main complications are reported as: breast stiffness, late seroma, lymph node enlargement, and silicone migration to distant organs. In addition, clinical manifestations related to autoimmune reactions have been reported, which highlight the silicone implant incompatibility syndrome (SIIS) (13).

However, gel bleeding is virtually ignored by the academy as a trigger point for these changes. Most articles advocate that gel bleeding is a rare event with no pathological significance. There are no studies in the literature demonstrating the frequency of this event in BMRI scans.

Recently we have described a new radiological finding, silicone-induced granuloma of breast implant capsule (SIGBIC) (14), which is defined as granulation tissue formed from a reaction between the breast implant fibrous capsule and free silicone corpuscle due to bleeding of the intact breast implant. We have described 3 BMRI features that are irrevocable for this diagnosis: 1. black-drop signal; 2. mass with hypersignal at T2-weighted sequences; and 3. late contrast enhancement (15). In addition, we described the mechanism of its intracapsular formation (16).

The aim of this study was to determine the ability of BMRI for predicting GB according to SIGBIC diagnosis with the 3 irrevocable features. Furthermore, we determined if the equivocal imaging features according to the latest BI-RADS lexicon could predict GB. This is the first prospective study to determine its prevalence in clinical practice.

## MATERIALS AND METHODS

This was a prospective observational study conducted at a single institution from March 2017 to August 2018. We evaluated patients referred for BMRI. All patients who had breast implants were included in the study. The exclusion criteria were patients with previous BMRI scan during the study period to avoid duplicity of the results, technically inappropriate scans, and scans without contrast injection. The study was conducted in accordance with local ethics committee regulations. Free informed consent was obtained from all patients.

We used a standard BMRI protocol for a 1.5-T imager (Magnetom Aera; Siemens Healthcare) with a dedicated eight-channel bilateral breast coil in the axial orientation. The acquired sequences were: 1. Axial T2-weighted fast spin-echo; 2. Sagittal Proton-density weighted; 3. Sagittal T2-weighted silicone selective that enhance the silicone signal; 4. Sagittal T2-weighted sequence with silicone suppression that extract the silicone signal; and 5. 4 sequential dynamic contrast sequences with fat suppression and reconstruction with subtraction.

BMRI images was read by 1 radiologist with 18 years of experience in BMRI. Nine BMRI findings were reported according to the BI-RADS lexicon, considered as equivocal signs:

1. Capsular contracture: when the anteroposterior diameter of the implant is increased with fibrous capsule thickening and contrast enhancement (Figures 1, 2, 3, and 4).
2. Intracapsular rupture: discontinuity of the breast implant surface, often characterized by: subcapsular line, keyhole sign, teardrop sign, and linguine sign restricted to intracapsular space.
3. Extracapsular rupture: discontinuity of the fibrous capsule with extravasation of the internal silicone contents.
4. Water droplets: foci of water signal inside the intact breast implant inferring loss of surface permeability (Figure 1).
5. Implant Rotation: displacement of the posterior surface of the breast implant, usually identified by the seal (Figure 2).
6. Extracapsular Siliconoma: extracapsular free silicone without signs of fibrous capsule rupture.
7. Enlarged intramammary lymph node (EILN): enlarged pericapsular intramammary lymph node (Figure 3). Two criteria were used to determine EILN: smaller diameter greater than 0.5cm and thickening of the cortical with reduced fatty hilum.
8. Pericapsular edema: non-mass pericapsular enhancement (Figure 4).
9. Intracapsular seroma: collection inside the fibrous capsule (Figures 1 and 2).

**Figure 1.**
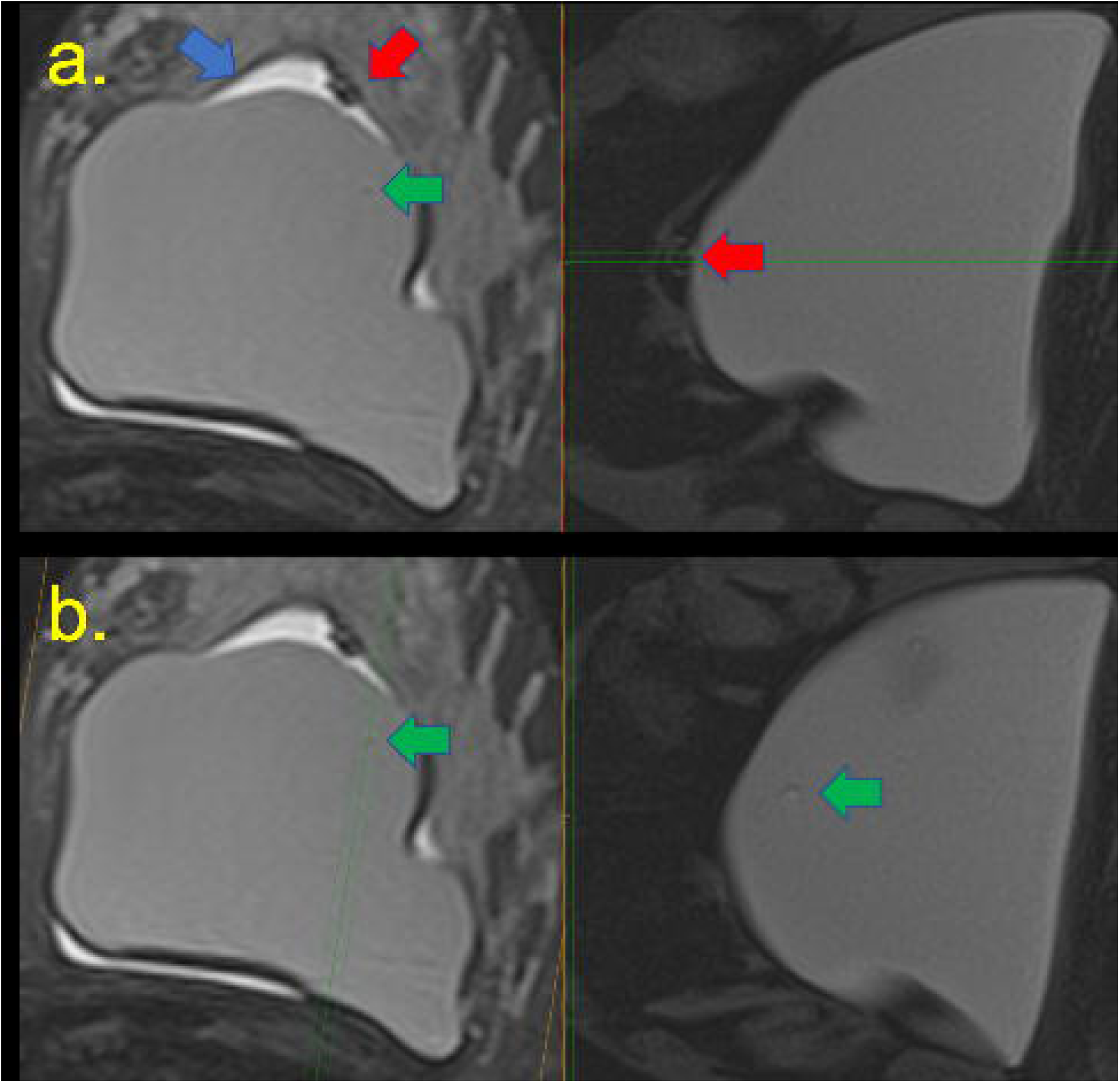
A 45-year-old woman 6 years after implant placement with left breast stiffness T2-weighted and silicone-sensitive images on the right and left, respectively. Blue and red arrows demonstrate an intracapsular seroma and mass that includes free silicone. (a) Intact breast implant. (b) Green arrow demonstrates water droplets.

**Figure 2.**
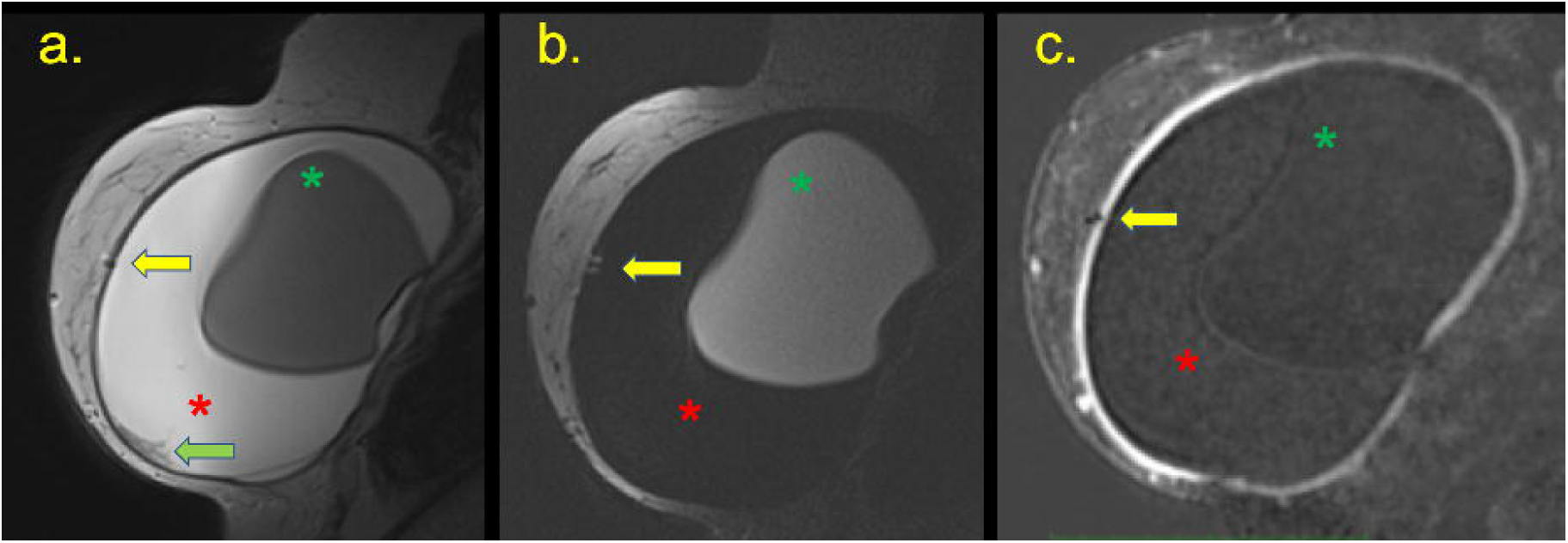
A 30-year-old woman who underwent an aesthetic procedure for silicone prosthesis four years ago, 1 week before an increase in volume and inflammation of the right breast (a) STIR sequencing presented a massive intracapsular collection (red asterisk) with intact and rotated breast implant (green asterisk), and free silicone in the fibrous capsule forming the granuloma (yellow arrow). (b) Identical results were found with T2W silicone-sensitive, and after the use of contrast. (c) Findings were compatible with the black drop signal. Intact breast implant.

**Figure 3.**
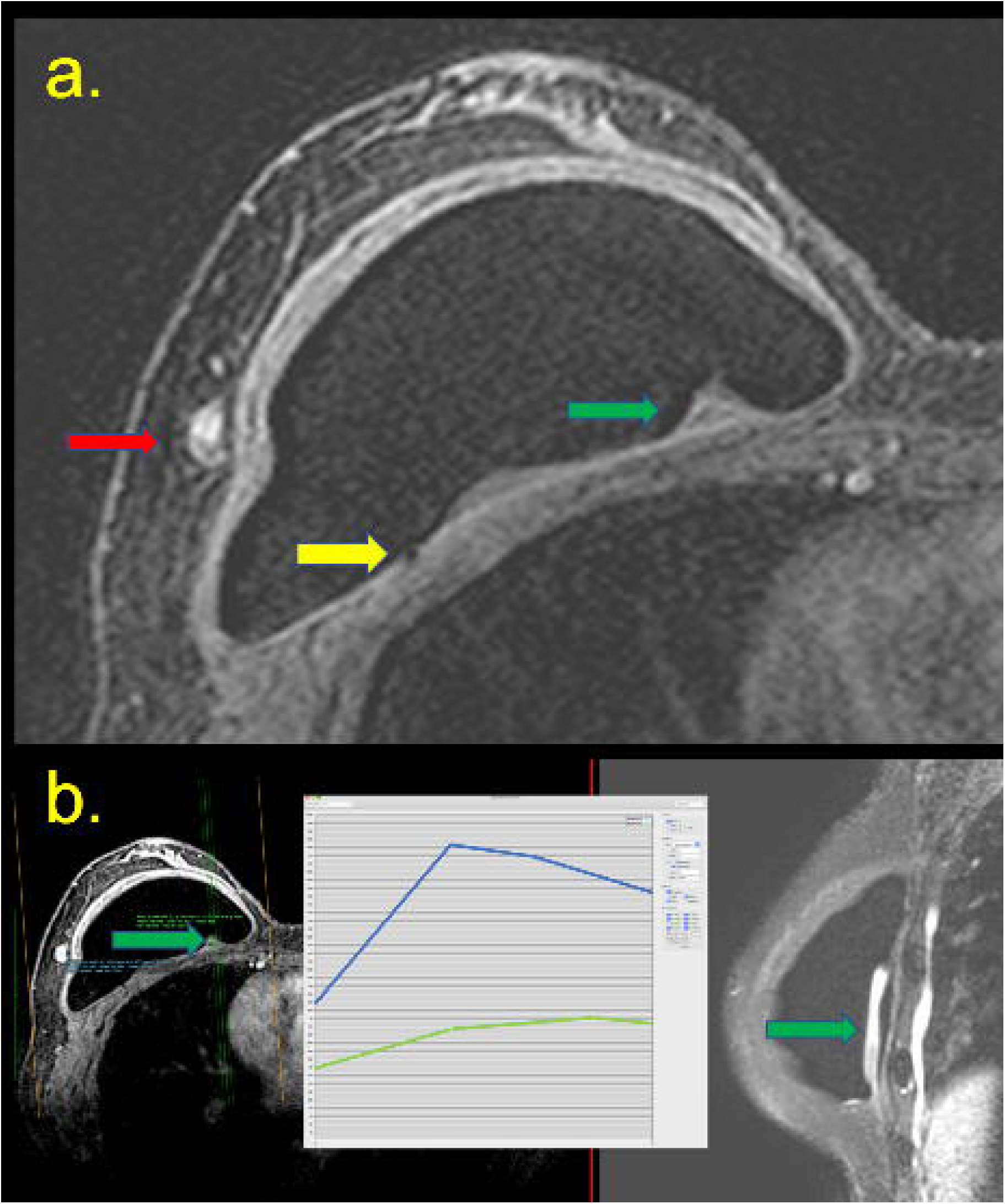
A 62-year-old patient underwent right breast repair surgery three years ago, followed by treatment with radiotherapy sessions (a) Arthralgia had been reported for 3 months. Intracapsular masses are shown in the pre-contrast sequence (green arrow), with enlarged intramammary lymph nodes (red arrow), and black-drop signal (yellow arrow). (b) A left mass was revealed by hypersignal on T2 imaging. (B). Intact breast implant.

**Figure 4.**
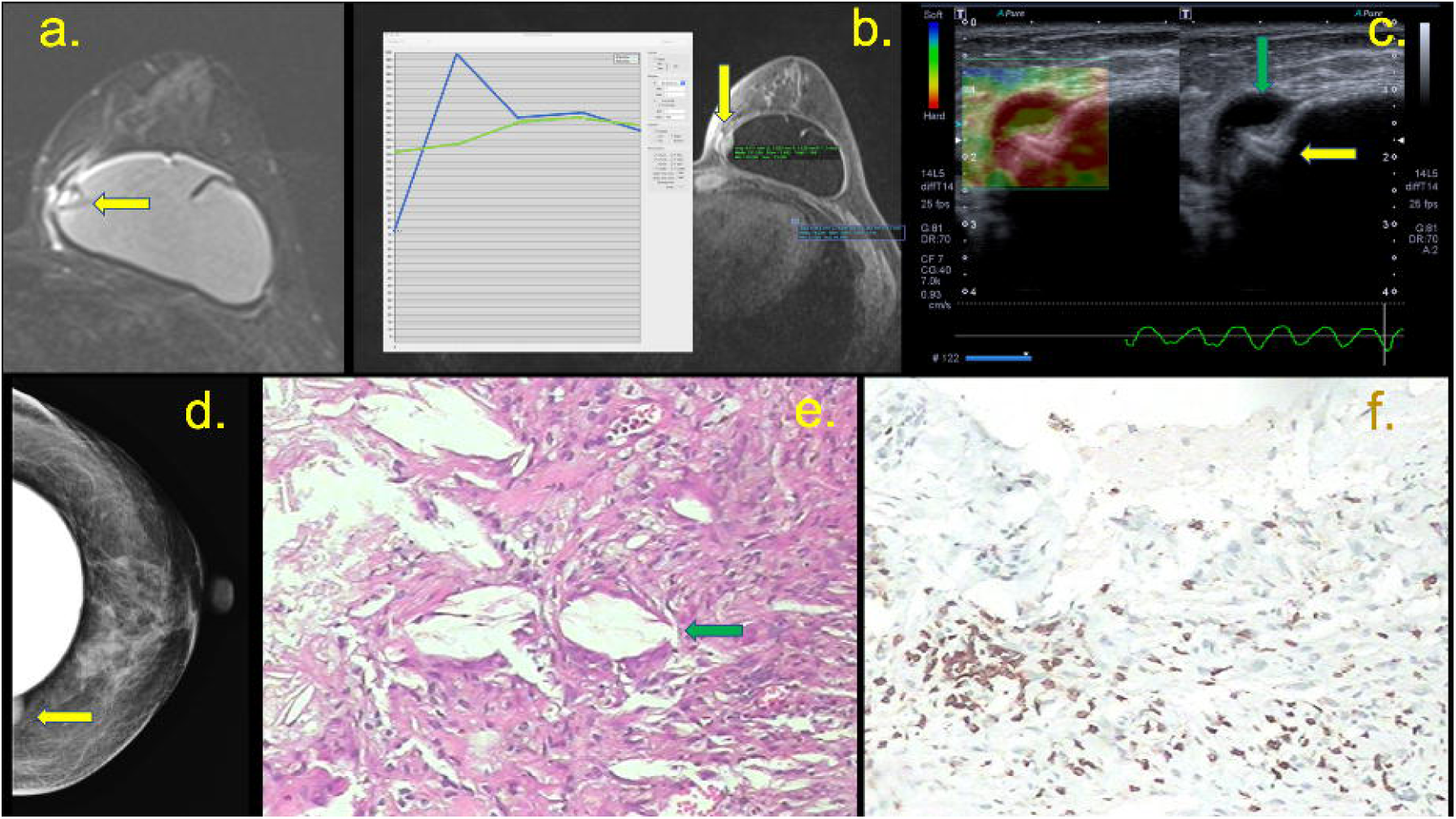
(a, b, c, d, e and f) A 37-year-old patient with a breast implant for 3 years, who showed a palpable mass in the medial quadrants of the left breast for 3 weeks (a) T2W fat-suppressed imaging showing a capsular contracture with a hypersignal mass in the medial quadrant and pericapsular edema (yellow arrow). The post-contrast sequence shows the late enhancement of this mass (green curve) compared with the heart (blue curve). (b) A pericapsular enhancement is shown (yellow arrow). (c) Ultrasound with elastography shows siliconoma at the site *via* BMRI, characterized by a hard mass at and snowstorm artifacts (yellow arrow), with a small collection at the periphery (green arrow). (d) Mammography shows a pericapsular mass in the medial quadrant (yellow arrow). (e) Histology showing free silicone particles and inflammatory cells (magnification, 50×). Immunohistochemistry for T lymphocyte (CD3) positive. Intact breast implant.

In addition to the features described by the BI-RADS lexicon, a further 3 new original features were incorporated that are reported with the presence of SIGBIC, considered as the tenth finding:

- 10.a. black drop signal: marked focus of hiposignal at T1 sequences in the fibrous capsule, silicone signal focus could be associated at silicone sensitive sequences, without enhancement in pos contrast sequences;
- 10.b. mass with hypersignal at T2 weighted sequence: intracapsular mass that could misdiagnose as seroma
- 10.c. late contrast enhancement: the mass (10b) shows enhancement at the late phase.

SIGBIC diagnosis was only determined when all the 3 irrevocable features were present.

Patients diagnosed with SIGBIC at BMRI underwent a second-look ultrasound scan using a device with 7.5–14 mHz multi-frequency probe (Aplio 300, Toshiba). All the patients who meet the SIGBIC BMRI diagnostic criteria underwent US-guided percutaneous core-biopsy or were directed for surgical capsulectomy. It was opted for the percutaneous biopsy in patients who have easy access for performing the procedures. In patients where the lesions were very deep or posterior to the breast implants, capsulectomy was opted. Percutaneous biopsy was performed using a 14-G needle attached to an automatic biopsy gun. At least 3 samples were collected. The remaining BMRI findings were followed up.

The specimens from biopsies or surgical procedures were evaluated in the same institution, where a diagnosis of SIGBIC was confirmed when the silicone particle was observed at microscopy, as described in a previous study. (Figure 5)

**Figure 5.**
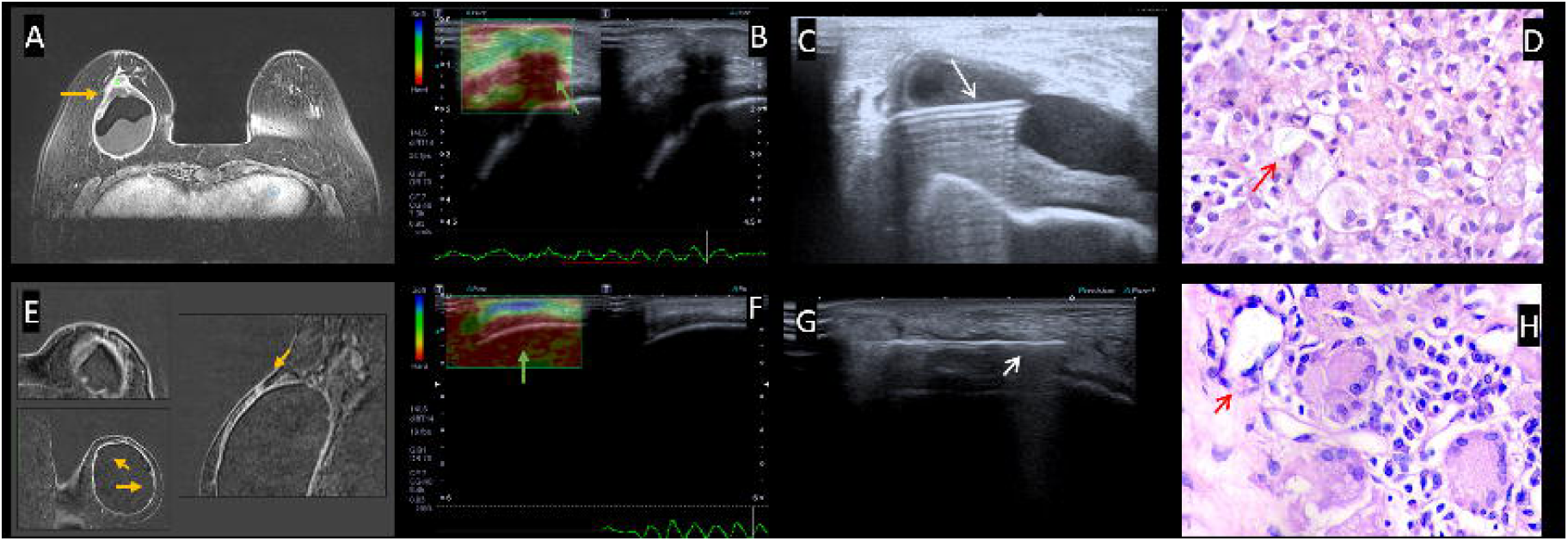
Examples of percutaneous breast biopsy in 2 patients. Patient 1 (A, B, C and B) and Patient 2 (e, f, g and h). At BMRI (A and E), findings suggestive of SIGBIC pointed by the yellow arrow. Ultrasound elastography (B and F) showing a hard mass at the fibrous capsule (green arrow). Biopsy of the mass (C and G) using first a fine needle aspiration to collect the intracapsular seroma (C) and core biopsy of the mass (G) represented by the white arrow. Histological specimens confirming silicone granuloma in red arrow.

Patients SIGBIC diagnosis were further divided into groups and compared to the SIGBIC findings, as follows:

- Type of surgery: aesthetic or reparative;
- Type of implant: silicone, saline, or double lumen;
- Location of the implant: retroglandular or retropectoral;
- Implant placement time.

The main objective of the study was to determine the ability of BMRI to predict gel bleeding in patients with SIGBIC diagnosis compared with histopathology as the gold standard. As secondary objectives we established what equivocal BMRI features could be related to GB. In this context, a univariate analysis was performed through Chi-Square tests (17) and Fisher’s Exact Test (18) and test of Mann-Whitney (19) for the case of categorical variables (20). The approach used was not the automatic Stepwise. Through the univariate analysis, the potential predictors for the response variable were selected, being considered a level of significance equal to 25% according to Kennedy and Bancroft criteria (21). The multivariate Logistic Regression model was only used for patients with histopathological proved SIGBIC in order to determine diagnosis ability using ONLY the stablished BI-RADS lexicon BMRI features.

We also evaluated, in order to verify if the adjusted models were adequate and if they had good predictive capacity, some measures of quality of adjustment using only the association of the main equivocal features for GB diagnosis: Pseudo R^2^ (22), AUC (area under the ROC curve), Sensitivity, Specificity, VPP, and the Hosmer-Lemeshow test (18). The software used in the analyzes was R (version 3.5.2).

For illustrative purposes, a descriptive analysis was also performed of the appropriate variables of interest. The absolute and relative frequencies were used. Whereas, for numerical variables description, positions, central tendency and dispersion were applied.

## RESULTS

We performed 2290 consecutive BMRI from March 2017 to August 2018. Of these, 736 patients had breast implants. Fifty-six patients were excluded from the study, 48 because they refused the injection of contrast medium, 6 because the presence of movement artifacts or difficulty in performing the silicone sequences, and 2 dues to previous BMRI scans during the study period. We also excluded 472 patients without the 3 diagnostic criteria for SIGBIC (equivocal features)

The remaining 208 patients with SIGBIC diagnosis by BMRI (irrevocable signs) were included in the study. All 208 patients with the irrevocable findings had GB confirmed by silicone corpuscles in histopathology. Histological specimens of 40 (19.9%) cases were obtained by surgical capsulectomy and 168 (80.1%) from percutaneous breast biopsy. There were no false positive results of GB adopting the SIGBIC irrevocable criteria. All capsular specimens showed silicone particles associated with inflammatory response at histology, and all patients submitted to surgical capsulectomy have no macroscopic signs of implant rupture.

### Univariate analysis

Table 1 shows the univariate analysis using Chi-Square test (17) for categorical explanatory variables with expected frequencies greater than 5 in all class and Fisher’s exact test (17) for the opposite cases. For the numerical variables (patient age and time of surgery), Mann-Whitney test analysis (20) was performed. The equivocal features with statistical significance selected in the univariate analysis (p<0.001) to predict GB diagnosis were: water droplet, enlarged intramammary lymph node, pericapsular edema, and intracapsular seroma. Type of surgery (p-value = 0.023), Implant type (p-value = 0.066) and patient age (p-value = 0.026) were also selected, once which had a p-value of less than 0.250. The variable capsular contracture did not participate in this analysis since it was observed in all patients.

**Table 1.** Univariate analysis of Breast MRI features.

| Variables | Category | SIGBIC (+) |  | P-value |
| --- | --- | --- | --- | --- |
|  |  | N (208) | % |  |
| Capsular Contracture | (-) | 0 | 0 | <0.001<sup>2</sup> |
|  | (+) | 208 | 30.6% |  |
| Intracapsular Rupture | (-) | 207 | 100.0% | - |
|  | (+) |  |  |  |
| Extracapsular Rupture | (-) | 208 | 30.8% | 0.319<sup>2</sup> |
|  | (+) | 0 | 0.00% |  |
| Water droplet | (-) | 142 | 25.7% | <0.001<sup>1</sup> |
|  | (+) | 66 | 51.97% |  |
| Implant Rotation | (-) | 198 | 31.2% | 0.274<sup>1</sup> |
|  | (+) | 10 | 22.2% |  |
| Siliconoma Extracapsular | (-) | 207 | 30.6% | 1.000<sup>2</sup> |
|  | (+) | 1 | 25.0% |  |
| Enlarged Intramammary Lymph Node (EILN) | (-) | 179 | 28.3% | <0.001<sup>1</sup> |
|  | (+) | 29 | 60.4% |  |
| Pericapsular edema | (-) | 179 | 27.9% | <0.001<sup>1</sup> |
|  | (+) | 29 | 74.4% |  |
| Intracapsular seroma | (-) | 166 | 27.7% | <0.001<sup>1</sup> |
|  | (+) | 42 | 51.2% |  |
| Indication | Screening | 11 | 9.8% | <0.001<sup>1</sup> |
|  | Diagnostic | 197 | 35.2% |  |
| Type of surgery | Aesthetic | 156 | 28.5% | 0.023<sup>1</sup> |
|  | Oncologic | 52 | 39.1% |  |
| Type of Implant | Silicone | 207 | 31.2% | 0.066<sup>2</sup> |
|  | Saline | 1 | 6.7% |  |
|  | Double | 0 | 0.0% |  |
| Location of Implant | Retroglandular | 141 | 29.2% | 0.290<sup>1</sup> |
|  | Retromuscular | 66 | 33.7% |  |
| Patient Age | Mean (S.E.) | 43.8 (0.8) |  | 0.026<sup>3</sup> |
| Time of surgery | Mean (S.E.) | 7.7 (0.5) |  | 0.878<sup>3</sup> |

<sup>1</sup>Chi-Square test. <sup>2</sup>Fisher's Exact test. <sup>3</sup>Mann-Whitney test
SIGBIC: silicone induced granuloma of breast implant capsule

### Multivariable analysis

A multivariate model of Logistic Regression (17) was adjusted from the variables selected in the univariate analysis and for this model the Backward method was applied for the final selection of the variables, considering a significance level of 5%.

Table 2 presents the final multivariate model for GB variable with the equivocal features described by the BI-RDADS lexicon. When positive an individual has the chance to have GB multiplied by 2.8 [1.8; 4.4] in the presence of water droplet, 3.07 [1.5; 6.1] in the presence of enlarged intramammary lymph node, 5.02 [2.3; 11.1] in the presence of pericapsular edema, and 2.4 [1.4; 4.1] for intracapsular seroma. Examination indication was significant (p-value <0.001) too, and considering the screening category as a reference, an individual with a diagnostic indication has a chance of having GB multiplied by 5.1 [2.6; 10.0].

**Table 2.**
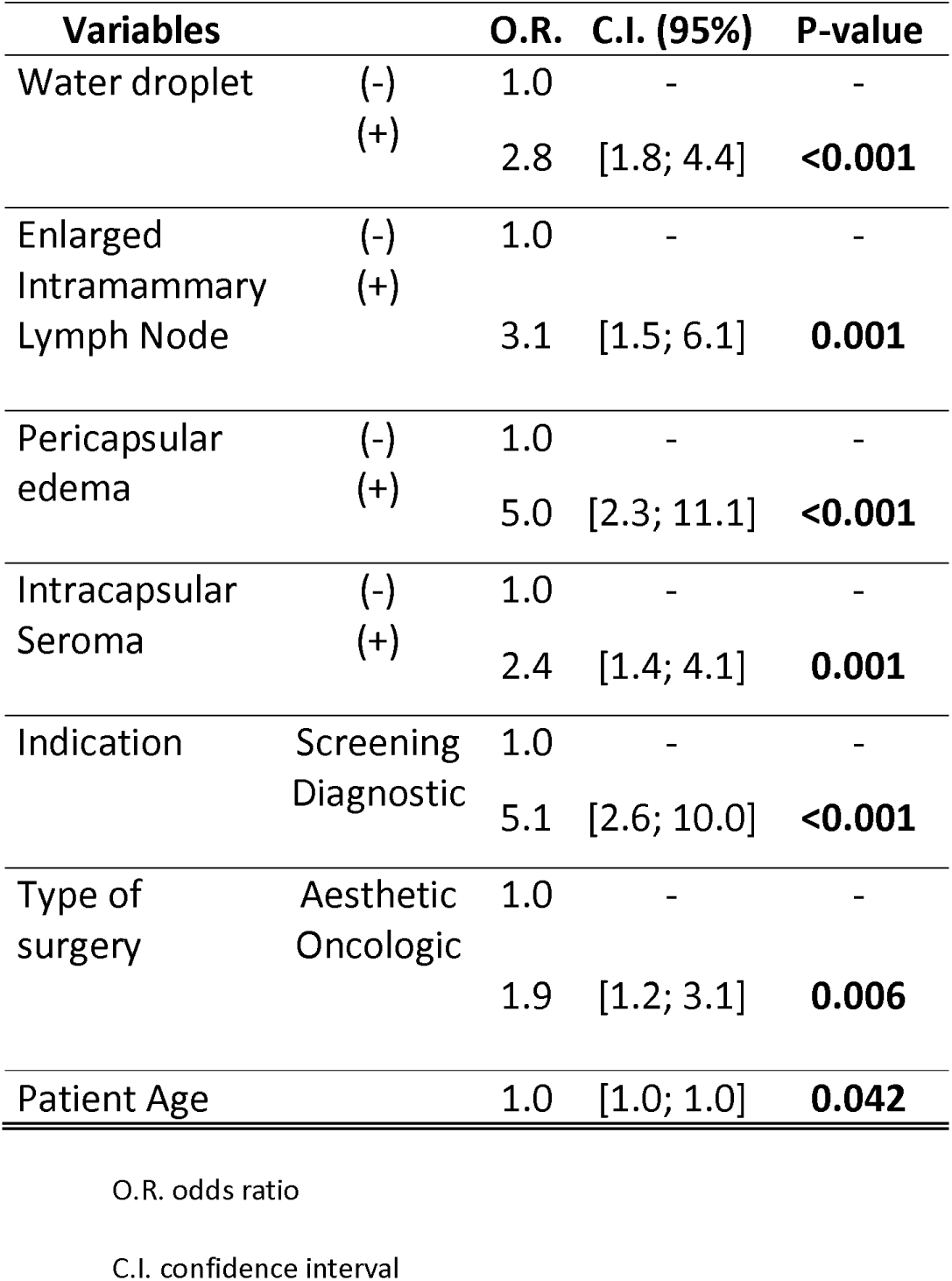
Multivariate analysis associating BMRI vaiables with and without gel bleeding

| **Variables** |  | **O.R.** | **C.I. (95%)** | **P-value** |
| --- | --- | --- | --- | --- |
| Water droplet | (-) | 1.0 | - | - |
|  | (+) | 2.8 | [1.8; 4.4] | **<0.001** |
| Enlarged<br>Intramammary<br>Lymph Node | (-) | 1.0 | - | - |
|  | (+) | 3.1 | [1.5; 6.1] | **0.001** |
| Pericapsular<br>edema | (-) | 1.0 | - | - |
|  | (+) | 5.0 | [2.3; 11.1] | **<0.001** |
| Intracapsular<br>Seroma | (-) | 1.0 | - | - |
|  | (+) | 2.4 | [1.4; 4.1] | **0.001** |
| Indication | Screening | 1.0 | - | - |
|  | Diagnostic | 5.1 | [2.6; 10.0] | **<0.001** |
| Type of<br>surgery | Aesthetic | 1.0 | - | - |
|  | Oncologic | 1.9 | [1.2; 3.1] | **0.006** |
| Patient Age |  | 1.0 | [1.0; 1.0] | **0.042** |

O. R. odds ratio
C.I. confidence interval

Table 3 shows the negative predictive value (NPV), Hosmer-Lemeshow test and Pseudo-R^2^ test for determination of GB for the final model adopting the most statistic significant equivocal BMRI features. It should be noted that: the area under the ROC curve (AUC) of the final model was 0.750 and the Pseudo-R2 was 24.79%. And the Hosmer-Lemeshow test for the final model indicated that the fit of the model was adequate (p-value = 0.795).

**Table 3.** Quality statistics of the multivariate model for GB diagnosis adopting water droplets, enlarged intramammary lymph nodes, pericapsular edema and intracapsular seroma.

| SEN | SPE | PPV | NPV | Accuracy | AUC | Hosmer-Lemeshow | Pseudo-R<sup>2</sup> |
| --- | --- | --- | --- | --- | --- | --- | --- |
| 0.740 | 0.638 | 0.474 | 0.848 | 0.669 | 0.750 | 0.795 | 24.79% |

SEN: sensitivity
SPE: specificity
PPV: positive predictive value
NPV: negative predictive value
AUC: area under curve

**Table 4.** Univariate analysis of Breast MRI features.

| Variables | Category | SIGBIC (-) |  | SIGBIC (+) |  | P-value |
| --- | --- | --- | --- | --- | --- | --- |
|  |  | N<br>(473) | % | N<br>(208) | % |  |
| Capsular Contracture | (-) | 123 | 26.1% | 0 | 0 | **<0.001**<sup>2</sup> |
|  | (+) | 349 | 73.9% | 208 | 30.6% |  |
| Intracapsular Rupture | (-) | 439 | 68.6% | 201 | 31.4% | 0.094<sup>1</sup> |
|  | (+) | 33 | 82.5% | 7 | 17.5% |  |
| Extracapsular Rupture | (-) | 468 | 69.2% | 208 | 30.8% | 0.319<sup>2</sup> |
|  | (+) | 4 | 100.00% | 0 | 0.00% |  |
| Water droplet | (-) | 411 | 74.3% | 142 | 25.7% | **<0.001**<sup>1</sup> |
|  | (+) | 61 | 48.0% | 66 | 51.97% |  |
| Implant Rotation | (-) | 437 | 68.8% | 198 | 31.2% | 0.274<sup>1</sup> |
|  | (+) | 35 | 77.8% | 10 | 22.2% |  |
| Siliconoma Extracapsular | (-) | 469 | 69.4% | 207 | 30.6% | 1.000<sup>2</sup> |
|  | (+) | 3 | 75.0% | 1 | 25.0% |  |
| Enlarged Intramammary Lymph Node (EILN) | (-) | 453 | 71.7% | 179 | 28.3% | **<0.001**<sup>1</sup> |
|  | (+) | 19 | 39.6% | 29 | 60.4% |  |
| Pericapsular edema | (-) | 462 | 72.1% | 179 | 27.9% | **<0.001**<sup>1</sup> |
|  | (+) | 10 | 25.6% | 29 | 74.4% |  |
| Intracapsular seroma | (-) | 432 | 72.2% | 166 | 27.7% | **<0.001**<sup>1</sup> |
|  | (+) | 40 | 48.8% | 42 | 51.2% |  |
| Indication | Screening | 109 | 90.8% | 11 | 9.8% | **<0.001**<sup>1</sup> |
|  | Diagnostic | 363 | 64.8% | 197 | 35.2% |  |
| Type of surgery | Aesthetic | 391 | 71.5% | 156 | 28.5% | **0.023**<sup>1</sup> |
|  | Oncologic | 81 | 60.9% | 52 | 39.1% |  |
| Type of Implant | Silicone | 456 | 68.8% | 207 | 31.2% | 0.066<sup>2</sup> |
|  | Saline | 14 | 93.3% | 1 | 6.7% |  |
|  | Double | 2 | 100.0% | 0 | 0.0% |  |
| Location of Implant | Retroglandular | 342 | 70.8% | 141 | 29.2% | 0.290<sup>1</sup> |
|  | Retromuscular | 130 | 66.3% | 66 | 33.7% |  |
| Patient Age | Mean (S.E.) | 41.6 (0.5) |  | 43.8 (0.8) |  | **0.026**<sup>3</sup> |
| Time of surgery | Mean (S.E.) | 7.3 (0.2) |  | 7.7 (0.5) |  | 0.878<sup>3</sup> |

<sup>1</sup>Chi-Square test. <sup>2</sup>Fisher's Exact test. <sup>3</sup>Mann-Whitney test
SIGBIC: silicone induced granuloma of breast implant capsule

### Descriptive analysis

It is observed that all patients with SIGBIC features had silicone corpuscles at histology and capsular contracture at BMRI. 18.7% had water droplet signs, 12.1% had intracapsular seroma and 7.1 had enlarged intramammary lymph node. The diagnostic indication for BMRI scan was prevalent (82.35%). The mean patient age value was 42.3 years with a standard deviation of 11.2 and the last surgery procedure was on average 7.4 years ago.

The mean time for patients diagnosed with SIGBIC was 7.7 years (median, 6 years). When evaluated if there is association between the diagnostic BMRI scan to the years passed after implant placement, the association to SIGBIC was 12 years (P = 0.0346) after surgical procedure for all patients. When we split these patients and assessed patients undergoing cosmetic surgery the association was 12 years (P=0.052) while that who underwent oncologic procedure was 7 years (P=0.0485). However, when evaluating implant location, we found no statistically significant difference in presence of SIGBIC between retroglandular or retromuscular implants.

## DISCUSSION

Currently, silicone bleeding is virtually ignored by academia as a possible trigger factor for complications related to silicone implants. Most articles illustrate silicone bleeding as rare event and of little clinical relevance. They justify this statement due to the evolution of breast implants, especially with the use of cohesive gel.

Since the description of SIGBIC by our group we observed an increasing prevalence of SIGBIC in our clinical practice; therefore, we conducted this prospective study to determine BMRI ability to predict GB in patients with SIGBIC diagnosis. We also intended to assess the association between BMRI equivocal features described by the latest BMRI BI-RADS lexicon and SIGBIC in predicting GB.

Reported breast implant-associated complications have recently been increasing, including BIA-ALCL and Breast Implant Illness in academia, mainstream and social media. Despite the reported severity associated with anaplastic lymphoma, the number of cases described in the literature remains small, with <650 cases reported, minimizing its relevance (11,12). Furthermore, the trigger point to develop this pathology remains unclear.

We recently described the radiological findings of a granuloma developed in the intracapsular compartment of breast implants that was formed by an immune response of the fibrous capsule to the free silicone particles bleeded from intact breast implants (16). All breast implants after a certain time or when subjected to stress situations (like heat and trauma) may alter the surface permeability may present gel bleeding of the internal contents or leakage of the shell (1). Both the silicone and saline implant have the substance polydimethilsilixonase (PDMS) as a shell component, and it is speculated that this substance can elicit an immune response when in contact with the fibrous capsule. Some patients have a higher risk for developing immune response, for example, those with SIIS. SIIS clinical manifestations may vary and depend on the intensity of the inflammatory reaction of the host. (16).

In our study, 30.6% patients with breast implants referred for BMRI scan at our facility fulfill the irrevocable BMRI criteria for SIGBIC diagnosis. All these cases were confirmed by histopathology by the presence of silicone corpuscles.

Due to the novelty of the study, we chose very restrictive criteria for the SIGBIC diagnosis. False positive results were not desired in this context. The black-drop signal consists of giant cell reaction to a foreign body in the fibrous capsule. The mass with hypersignal at T2-weighted sequence is the development of granulation tissue in the contacted area between the silicone corpuscle and the fibrous capsule. Finally, the late contrast enhancement is because of the poor intracapsular vascularization due to the fibrous capsule protective barrier and could differentiate the granuloma from intracapsular seroma. When the three criteria were met, we could predict GB in all cases.

We found statistical significance association between GB and some of the equivocal BMRI BI-RADS lexicon features: capsular contracture, water droplets, enlarged intramammary lymph node, pericapsular edema, and intracapsular seroma. All probably due to leakage of silicone gel in intact implants. When used the Hosmer-Lemeshow test and pseudo-R2 to associate the presence of GB with only the equivocal BMRI features, there was found good sensitivity, specificity and diagnostic accuracy for predicting GB. These findings could support the hypothesis of GB be underdiagnosed in our clinical practice where most of the articles states that is a rare event.

We hypothesized that water droplets are related to permeability loss in the breast implant shell. When intracapsular fluid enters through the implant shell, a chemical reaction occurs with the inside silicone content or the silicone-made surface of the implant, which leads to silicone residues into the intracapsular compartment. Water-droplet corresponds to a macroscopic diagnosis, where the findings are seen with the naked eye by BMRI. It is speculated that the water-droplets diagnosis by BMRI may be underestimated if compared with microscopy. (Figure 6) These findings were similar to that described as: *“breast implants, from clear to cloudy”* (23) where the authors report color changes of intact breast implants due to a chemical reaction with intracapsular fluid. In our study, when water droplets are present the risk of SIGBIC is increased in 2.82.

**Figure 6.**
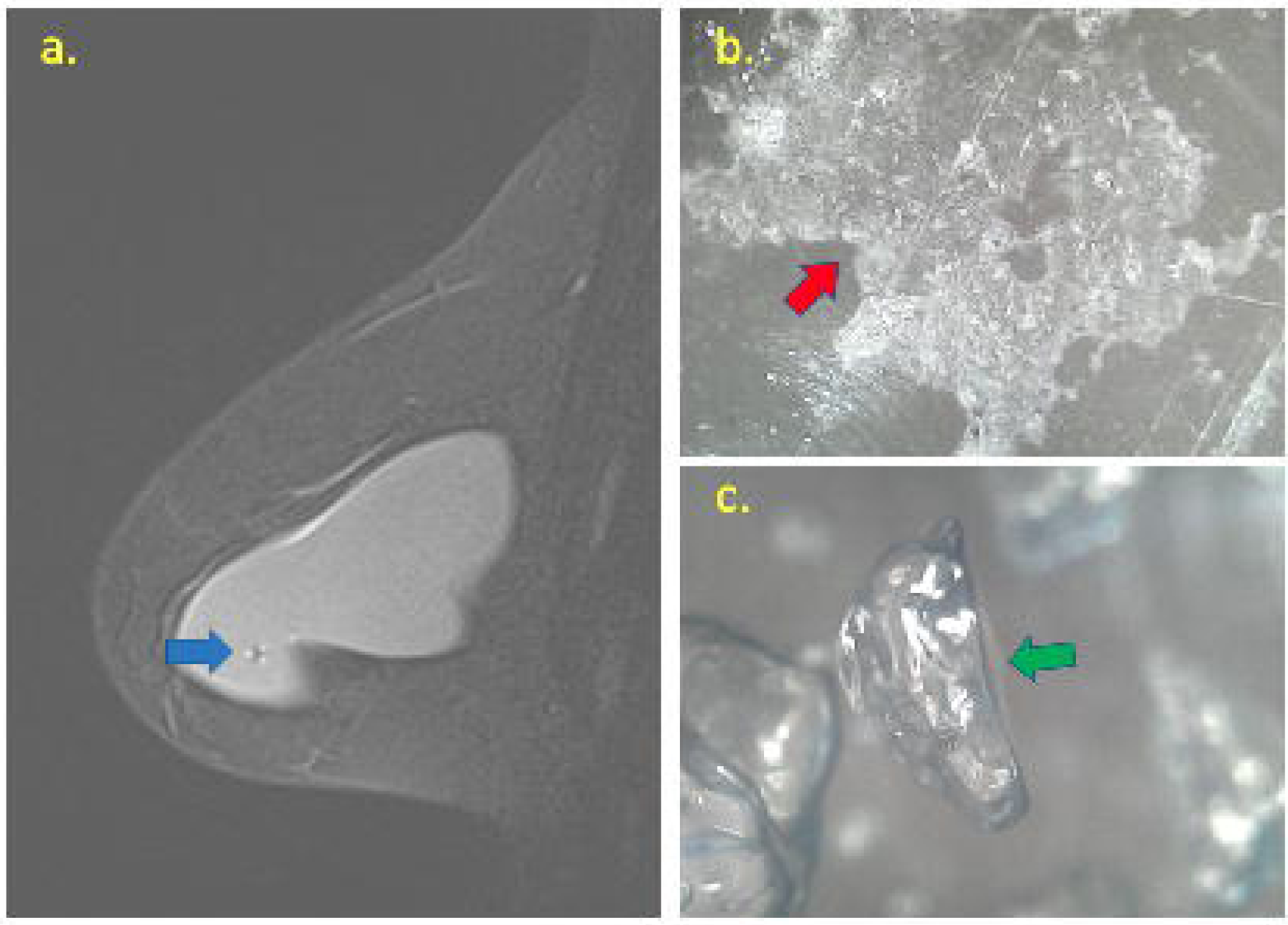
Correlation between water droplets at BMRI and microscopy of implant shell. At BMRI (a) is possible to see a focal signal change inside the breast implant pointed by the blue arrow. At the microscopy evaluation of implant shell (b) is possible to note silicone leakage pointed by the red arrow. Inside the implant with a 1.600-fold microscopy (c) it is possible to see the water droplets formed from a chemical reaction between the intracapsular fluid and the silicone gel content.

There was also a correlation between surgery type and GB. Oncologic surgery time and SIGBIC diagnosis up to 7 years after surgery, which may be due to greater manipulation of the implant in therapeutic surgery, increased surgical times, and radiotherapy sessions (28).

We observed a high prevalence of GB in our study. Interestingly, no previous study has reported this occurrence neither the relation of GB with BMRI findings. There may be different reasons for this. First, culturally most BMRI protocols for breast implant evaluation are performed without the use of contrast media. This factor may lead to a misdiagnosis of SIGBIC as late seroma. Second, there may be a lack diagnostic expertise. For example, before 2017, our service had not reported a positive diagnosis of SIGBIC. However, some patients were diagnosed with SIGBIC during the study period, following persistent clinical complaints, and when comparing to previous exams; SIGBIC was retrospectively diagnosed.

The relevance of this manuscript is the novelty of the issue and to alert to the possibility of gel bleeding as a precursor to complications inherent to silicone breast implants. The results exhibit the high frequency of gel bleeding in this study, where it describes the restricted criteria for diagnosis. Currently, many diseases are related to silicone implants, such as BIA-ALCL, SIIS, and Breast Implant Illness (BII), without a clear hypothesis of what is the trigger point for its development. With the information obtained from the BMRI, the doctor can more accurately manage patients with complaints related to breast implants.

This study has limitations. First, we only evaluated diagnostic BMRI in a single center. Future prospective multicenter studies should be conducted to confirm our findings. We are initiating a multicenter study involving three different breast diagnostic centers to validate our results. An additional limiting factor was that only patients diagnosed with SIGBIC underwent percutaneous biopsies or surgical capsulectomies. As an observational study, it was not accepted by our ethical committee to have a control group submitted to breast biopsy. However, we were able to illustrate that MRI can diagnose gel bleeding, the novelty of this study. Future studies should access the histology of patients with capsular contracture to search for free silicone in the fibrous capsule. It is important to emphasize the importance of pathologist and radiologist training to diagnose SIGBIC.

Our study supports that GB prediction could be performed adopting the 3 irrevocable SIGBIC criteria proposed by the authors. It also demonstrates that equivocal BMRI features related to GB were capsular contracture, water droplets, enlarged intramammary lymph node, pericapsular edema and intracapsular seroma. Based on our findings, we suppose it is underdiagnosed in clinical practice and could explain most of the implant reported complications.

## Data Availability

all data is available in authors files.

## List of abbreviations

SIGBIC: silicone-induced granuloma of breast implant capsule
BMRI: breast magnetic resonance imaging
GB: gel bleeding
BIA-ALCL: breast implant-associated anaplastic large cell lymphoma
EILN: enlarged intramammary lymph node
SIIS: silicone implant incompatibility syndrome

## References

1. Kappel R, Boer L (2016) Gel Bleed and Rupture of Silicone Breast Implants Investigated by Light-, Electron Microscopy and Energy Dispersive X-ray Analysis of Internal Organs and Nervous Tissue. Clin Med Rev Case Reports 3:1–9.

2. Willem J, Tervaert C (2018) Autoinflammatory/autoimmunity syndrome induced by adjuvants (ASIA; Shoenfeld’s syndrome): A new flame. Autoimmun Rev 17:1259–1264.

3. Report C (2018) Hepatic Infiltration by Silicone in a Patient With ASIA Syndrome. 67:2017–2108.

4. Malahias M, Jordan DJ, Hughes LC, Hindocha S, Juma A (2016) A literature review and summary of capsular contracture: An ongoing challenge to breast surgeons and their patients. Int J Surg Open 3:1–7.

5. Leberfinger AN, Behar BJ, Williams NC, et al (2017) Breast implant-associated anaplastic large cell lymphoma: A systematic review. JAMA Surg 152:1161–1168.

6. Pastorello RG, D’Almeida Costa F, Osório Cabt,et al (2018) Breast implant-associated anaplastic large cell lymphoma in a Li-FRAUMENI patient: a case report. Diagn Pathol 13:10.

7. Ezekwudo DE, Ifabiyi T, Gbadamosi B, Haberichter K, et al (2017) Case Report Breast Implant–Associated Anaplastic Large Cell Lymphoma: A Case Report and Review of the Literature. Case Rep Oncol Med 6478467

8. Orofino N, Guidotti F, Cattaneo D, et al (2016) Marked eosinophilia as initial presentation of breast implant-associated anaplastic large cell lymphoma. Leuk Lymphoma 57:2712–2715.

9. De Boer M, Van Der Sluis WB, De Boer JP, et al (2017) Breast implant-associated anaplastic large-cell lymphoma in a transgender woman. Aesthetic Surg J 37:NP83–NP87.

10. Hart AM, Lechowicz MJ, Peters KK, Holden J, Carlson GW (2014) Breast implant-associated anaplastic large cell lymphoma: Report of 2 cases and review of the literature. Aesthetic Surg J 34:884–894.

11. Administration USF and D. (2018) Breast Implant-Associated Anaplastic Large Cell Lymphoma (BIA-ALCL). Available from: https://www.fda.gov/MedicalDevices/ProductsandMedicalProcedures/ImplantsandProsthetics/BreastImplants/ucm239995.htm. Accessed 20 Jun 2019

12. Coroneos CJ, Selber JC, Ii ACO, Butler CE, Clemens MW (2019) US FDA Breast Implant Postapproval Studies Long-term Outcomes in 99,993 Patients. 259:30–36.

13. Watad A, Rosenberg V, Tiosano S, et al (2018) Silicone breast implants and the risk of autoimmune/rheumatic disorders: a real-world analysis. Int J Epidemiol 47(6):1846–1854.

14. Fleury E de FC, Rêgo MM, Ramalho LC, et al (2017) Silicone-induced granuloma of breast implant capsule (SIGBIC): similarities and differences with anaplastic large cell lymphoma (ALCL) and their differential diagnosis. Breast Cancer 9:133–140.

15. de Faria Castro Fleury E, Gianini AC, Ayres V, Ramalho LC, Seleti RO, Roveda D (2017) Breast magnetic resonance imaging: tips for the diagnosis of silicone-induced granuloma of a breast implant capsule (SIGBIC). Insights Imaging. 8:439–446.

16. de Faria Castro Fleury E, D’Alessandro GS, Lordelo Wludarski SC (2018) Silicone-Induced Granuloma of Breast Implant Capsule (SIGBIC): Histopathology and Radiological Correlation. J Immunol Res 6784971.

17. AGRESTI, A (2002) Categorical analysis. New York: John Wiley.

18. Agresti, A.; Kateri, M (2011) Categorical Data Analysis. Gainesville, Florida: John Wiley, v. 45.

19. Efroymson, M. A (1960) Multiple regression analysis. In: Mathematical methods for digital computers. New York, N.Y.: John Wiley, 191–203.

20. Hollander, M.; WOLFE D. A (1999) Nonparametric Statistical Methods. 2nd. ed. New York, N.Y.: John Wiley & Sons.

21. Kennedy WJ, Bancroft, T (1971) Model Building for Prediction in Regression Based Upon Repeated significance Tests. Ann. Math. Statist 4:1273–1284 doi:10.1214/aoms/1177693240.

22. Nagelkerke, N. J. D (1991) A note on a general definition of the coefficient of determination. Biometrika, 1991.

23. Taylor EM, Sackeyfio R, Grant RT (2014) Clear to Cloudy: Silicone Breast Implants In Vivo. Aesthetic Plast Surg 38:827–829.

24. Schubert DW, von Hanstein H, Daenicke J, Werner S, Horch RE (2018) Compressive and cyclic loading of silicone breast implants and their effect on shape resilience and reliability of the shell material. Polym Int. 67:380–385.

